# Improving Automated Diagnosis of Middle and Inner Ear Pathologies by Estimating Middle Ear Input Impedance from Wideband Tympanometry

**DOI:** 10.64898/2026.03.26.26349034

**Authors:** Anna Frazier Kamau, Gabrielle R. Merchant, Hideko Heidi Nakajima, Stephen T. Neely

## Abstract

Conductive hearing loss (CHL) with a normal otoscopic exam can be difficult to diagnose because routine clinical measures such as audiometric air-bone gaps (ABGs) can identify a conductive component but often cannot distinguish among specific underlying mechanical pathologies (e.g., stapes fixation versus superior canal dehiscence, which may produce similar audiograms). Wideband tympanometry (WBT) is a fast, noninvasive test that can provide additional mechanical information across a broad range of frequencies (200 Hz to 8 kHz). However, WBT metrics are influenced by variations in ear canal geometry and probe placement and can be challenging to interpret clinically. In this study, we extend prior WBT absorbance-based classification work by estimating the middle ear input impedance at the tympanic membrane (Z_ME_), a WBT-derived metric intended to reduce ear canal effects. To estimate Z_ME_, we fit an analog circuit model of the ear canal, middle ear, and inner ear to raw WBT data collected at tympanometric peak pressure (TPP). Data from 27 normal ears, 32 ears with superior canal dehiscence, and 38 ears with stapes fixation were analyzed. A multinomial logistic regression classifier was trained using principal component analysis (retaining 90% variance) and stratified 5-fold cross-validation with regularization. We compared feature sets based on ABGs alone, ABGs combined with absorbance, and ABGs combined with the magnitude of Z_ME_. The combination of ABGs and the magnitude of Z_ME_ produced the best performance, achieving an overall accuracy of 85.6% compared to 80.4% for ABGs alone and 78.4% for ABGs combined with absorbance. These results suggest that incorporating model-derived middle ear impedance features with standard audiometric measures (ABGs) can improve automated pathology classification for stapes fixation and superior canal dehiscence.

## INTRODUCTION

### Difficulties with Diagnosis

Conductive hearing loss (**CHL**) is caused by a mechanical pathology that results in a disruption of sound transmission from the outer to the inner ear. Because the sensorineural system is functioning, CHL has the potential to be treated, for example with a surgical solution or bone-conduction hearing aid. However, diagnosing the underlying cause of middle or inner ear disorders that result in CHL can be challenging and despite technological advances, diagnostic approaches have remained largely unchanged for decades. Initial diagnostic efforts often include performing audiometric testing to measure the patient’s air-bone gaps (**ABGs**). Though such measures may inform of a CHL, they do not provide information regarding the specific pathology responsible for the hearing loss. For example, ABGs can be similar for a middle ear lesion such as stapes fixation (**SF**) and an inner ear lesion such as superior canal dehiscence (**SCD**) (Merchant et al. 2007). ABGs are only tested up to 4 kHz, so hearing loss above this frequency is typically presumed to be sensorineural; however, loosening of the ossicular chain can cause high-frequency CHL (Farahmand et al. 2016), thus can be diagnostically missed. Standard tympanometry at 226 Hz is often measured to diagnose some mechanical pathologies such as middle ear effusion or tympanic membrane perforation, but is unable to distinguish middle and inner ear pathologies (Margolis et al. 1999; Nakajima et al. 2005). Consequently, if the otoscopic exam appears normal, it is difficult to know the pathology responsible for the hearing loss.

Without a good initial diagnosis of the pathology underlying a hearing loss, clinicians have limited information in choosing the next appropriate diagnostic test, referral, or possible treatment. Further diagnostic testing may include high-resolution temporal bone computed tomography (**CT**) with radiation exposure, specialized measurements such as vestibular evoked myogenic potential (**VEMP**) which may only be available in specialized centers, and/or surgical middle ear exploration which is invasive and still may not always determine the pathology. Without a reliable working diagnosis of the underlying pathology at initial assessment, patients with CHL may experience delayed or inappropriate treatment, undergo unnecessary procedures, accrue increased healthcare costs, and ultimately experience reduced quality of care.

### Wideband Tympanometry

As described above, standard tympanometry is commonly measured by audiologists to provide some information about the mechanics of the ear, such as tympanic membrane perforation and effusion. Standard tympanometry presents positive and negative static pressure in the ear canal while also presenting a pure tone (most commonly at 226 Hz). The 226 Hz sound reflected from the tympanic membrane is then measured. A similar measure, wideband tympanometry (**WBT**), is as convenient to perform as standard tympanometry, but can provide more information sensitive to pathologies of the ossicles and inner ear. As in standard tympanometry, WBT also presents static pressures in the ear canal, but WBT simultaneously presents a wider range of acoustic frequencies beyond just 226 Hz, generally ranging from 200 to 8000 Hz. Therefore, WBT provides more comprehensive information about the mechanical characteristics of the ear, such as complex mass, stiffness, and damping. WBT measures are clinically informative due to their sensitivity to pathologies affecting middle ear and inner ear mechanics (Allen et al. 2005; Merchant and Neely 2021). However, WBT data is challenging to interpret for clinicians and therefore has not been adopted for initial diagnostic workup at most clinics, and the type of data and utilization of the data varies among clinicians (Al-Salim et al. 2024).

Because of the difficulty in interpreting the complex and abundant WBT data, algorithms, especially with machine learning tools, can automate the process and provide a richer understanding of ear mechanics. Towards this goal, a recent study from our group by Eberhard et al. (2024) showed that a multinomial logistic regression algorithm could produce good diagnostic estimates for two mechanical ear pathologies: SCD and SF, two very different pathologies that can have similar audiograms. The algorithm incorporated absorbance, a derivative of WBT (Feeney et al. 2013), in addition to ABGs, and achieved very good accuracy for differentiating pathologies.

### Limitations with Absorbance and Ear Canal Impedance

Although WBT can be a valuable diagnostic tool, several factors can introduce variability in WBT measurements. The WBT probe (with speaker output and microphone tube tip) is inserted to a stable, leak-free depth, but its exact position within the ear canal varies across tests and across individuals. Therefore, the measured acoustic impedance of the air between the probe tip and the tympanic membrane (i.e., the ear canal impedance), varies due to probe placement and ear canal length.

Although absorbance (with only magnitude information) is generally assumed to be insensitive to probe placement, this assumption is based on an idealized model of a rigid-walled tube with a constant cross-sectional area (Rosowski et al. 2013). In practice, ear canal size and shape vary significantly across subjects (Balouch et al. 2023; Voss et al. 2025). Moreover, even under ideal conditions, absorbance varies with cross-sectional area because it is computed using the characteristic impedance, which itself depends on that area. Therefore, both anatomical variability and probe placement differences can meaningfully affect measured absorbance.

### Purpose of our Study

Given these limitations associated with absorbance measurements, a WBT-derived metric that *removes the effect of the <u>ear canal</u> impedance* may offer improved diagnostic value for CHL. Specifically, estimating the input impedance at the *<u>middle ear</u>* (*Z*_*ME*_, **Figure 1**), that is, the impedance from the ear canal just in front of the tympanic membrane (thus including the middle and inner ear), may better represent the mechano-acoustic effects of pathologies without the varying anatomical and acoustic effects of the ear canal. We hypothesize that by removing ear canal effects, this middle ear impedance-based approach will improve the diagnostic capabilities of our multinomial logistic regression algorithm for diagnosing middle and inner ear pathologies.

**Figure 1.**
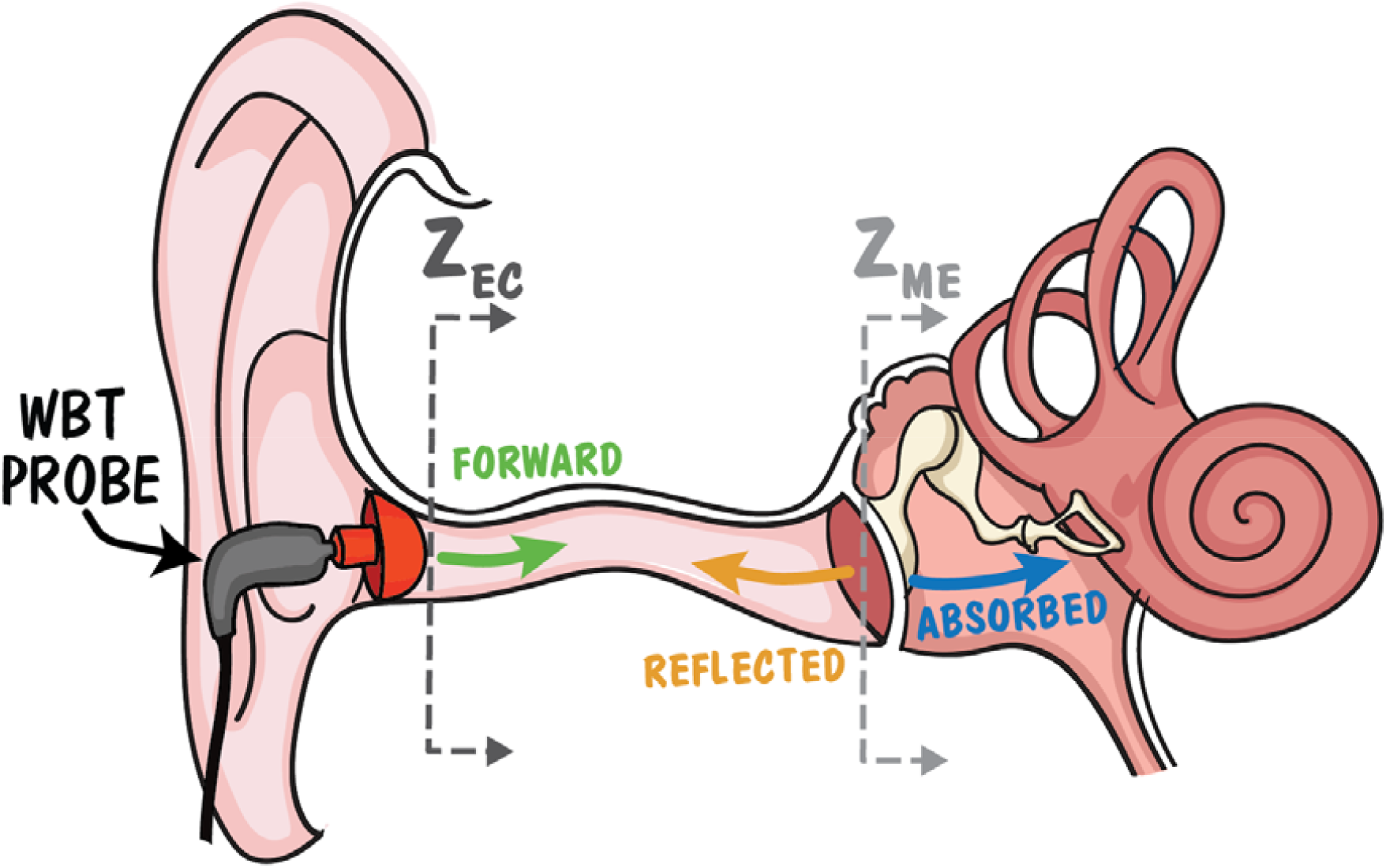
Illustration of the WBT measurement with arrows representing forward sound pressure delivered from the probe microphone (green), reverse sound pressure measured by the probe microphone (yellow), and the sound energy absorbed by the ear (blue). Dashed lines are used to indicate the planes of Z_EC_ and Z_ME_. Image adapted from Adobe Stock image #271603767.

The objectives of the present study were to advance our earlier strategy (Eberhard et al. 2024) by combining middle ear modeling principles with an estimate of *Z*_*ME*_ (instead of absorbance), and determine if it could (1) provide insights regarding the mechano-acoustic effects of SCD and SF and (2) offer additional diagnostic benefits compared to combining ABGs and absorbance.

## MATERIALS AND METHODS

### Subjects

This study analyzed data from 27 normal ears, 32 ears with SCD, and 38 ears with SF. The 70 pathological ears with SCD and SF included here were also used in our previous study (Eberhard et al. 2024). Pathological diagnosis was confirmed via surgery for SF and high-resolution CT scan showing a clear dehiscence (not thin bone) for SCD.

All ears had no history of ear surgery, no evidence of concurrent abnormalities (otitis media, cholesteatoma, or both SF and SCD), and a normal otoscopic exam prior to data collection. Additionally, normal ears had air and bone conduction thresholds ≤15 dB HL from 0.25 to 4 kHz, and air conduction thresholds ≤20 dB HL from 6 to 8 kHz. Standard 226 Hz tympanograms were considered normal if they met the following criteria: pressure between -100 to +50 daPa, admittance between 0.3 to 1.8 mmho, and ear canal volume between 0.3 to 2.5 mL (Gelfand & Calandruccio 2023). These measurements were performed at Massachusetts Eye and Ear. The study was approved by the Institutional Review Board at Massachusetts General Brigham (IRB Protocol #2019P003517).

### Measurement Procedure

WBT measurements were performed using the commercially available Interacoustics Titan WBT system. Instead of the standard clinical software, we used the Titan Research Platform to obtain the “raw” WBT measurements. These raw WBT measurements were the sound pressures measured by the microphone without any manipulation to the data, such as smoothing or noise-reduction (which occur within the Titan Suite Clinical Platform). To collect measurements, we used a custom user interface developed with MATLAB at Boys Town National Research Hospital (Merchant et al. 2021; Eberhard et al. 2022). Thévenin calibration of the system was conducted at the start of each test day (Nørgaard et al. 2017). During each measurement, a train of identical wideband click sound stimuli (0.2 to 8 kHz) was presented while the static pressure within the ear canal was varied between 200 and -300 daPa. After each measurement, we removed artifacts caused by measurement noise using our signal processing techniques described in Eberhard et al. (2022).

Participants underwent audiometric testing as part of their clinical diagnostic workup. Audiometric data were retrieved from medical records or research databases. All audiometry was performed within six months of the WBT measurement, with the majority tested on the same day. For each ear, the ABGs were computed as the differences between the air and bone conduction threshold at each frequency (0.25 to 4 kHz).

### WBT Measurements & Calculations

The Titan device presents a train of clicks (0.2-8 kHz) while the ear canal static pressure is downswept. The device measures the reflected pressure in the ear canal via a microphone (**Figure 1**). For this study, we restricted our analysis to measurements obtained at the tympanic peak pressure (TPP), defined as the static ear canal pressure at which the average absorbance between 0.38 and 2 kHz is maximized. Using measured pressure and the Thévenin calibration, we calculate the input impedance at the plane of the probe tip, *Z*_*EC*_ (Merchant 2014). *Z*_*EC*_ is the impedance from the tip of the probe that is inserted into the ear canal and includes the impedance of the rest of the ear canal to the tympanic membrane, the middle ear, and the inner ear (**Figure 1**).

We then calculate reflectance (*Γ*), which is the complex ratio of the reverse pressure to forward pressure (**Figure 1**). *Γ* can also be expressed in terms of input impedance *Z*_*EC*_ and characteristic impedance of the ear canal *Z*_*0*_,

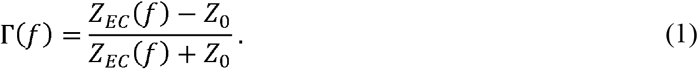

The characteristic impedance of the ear canal *Z*_*0*_ is estimated as,

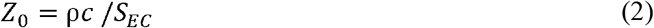

where the ear canal is assumed to be a tube with constant cross-sectional area *S*_*EC*_= 44.18 mm (diameter of 7.5 mm), ρ is the density of air, and *c* is the speed of sound in air.

From reflectance *Γ*, we can derive absorbance (𝔸) which varies by frequency and is related to the absorbed energy behind the tympanic membrane:

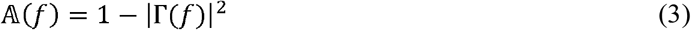

𝔸is the WBT-derived measurement often used in previous publications (Feeney et al. 2013; Eberhard et al. 2024) and contains the magnitude but not the phase information in the WBT data.

### Computational Model of the Ear

*Z*_*EC*_ includes the acoustic influence of the ear canal, middle ear, and inner ear (**Figure 1**). As previously mentioned, the acoustic influence of the ear canal can vary across ears due to anatomical variations and probe placement. To classify normal and pathological ears, with conditions related to the middle and inner ear, we want to remove the varied acoustic influences of the ear canal. To do so, we used the computational model (Merchant and Neely 2023) shown in **Figure 2**. *Z*_*M E*_ represents the input impedance starting from the end of the ear canal, just in front of the tympanic membrane, including the middle and inner ear. *Z*_*EC*_ includes the influence of the ear canal with non-uniform areas from the end of the probe tip up to the tympanic membrane, and the middle and inner ear components.

**Figure 2.**
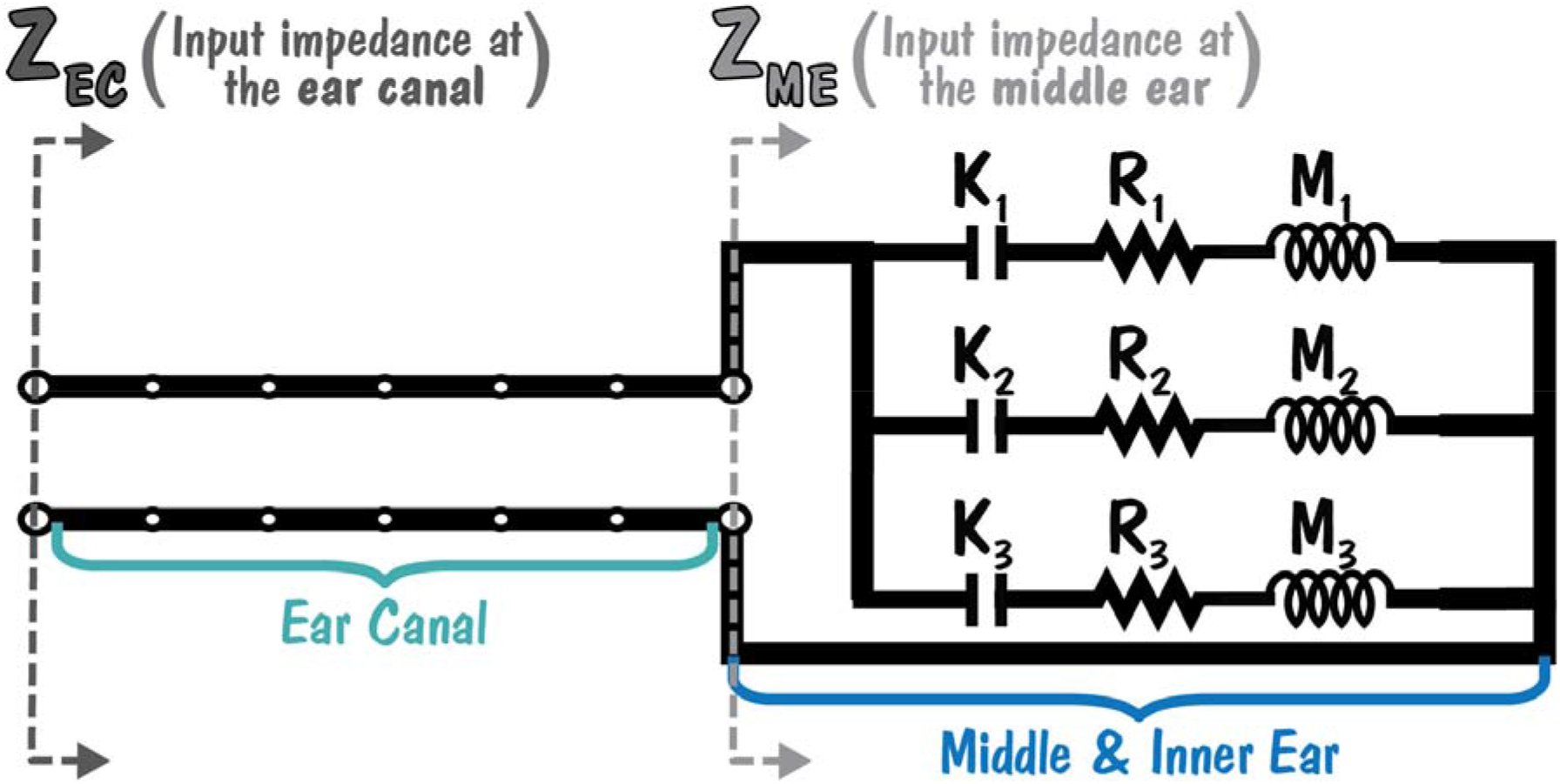
Analog circuit model of the input impedance, Z_EC_, starting at the tip of the probe and influenced by the ear canal and middle and inner ear. The ear canal is represented as a nonuniform transmission line composed of six concatenated cone segments. The input impedance Z_ME_ starting just in front of the tympanic membrane is modeled as three parallel branches, each containing stiffness K, damping R, and mass M components representing the influence of both the middle and inner ear.

The ear canal is modeled as a nonuniform transmission line composed of six concatenated truncated-cone sections. It is numerically implemented as a lossless two-port ABCD transmission matrix (Lewis and Neely, 2015; Merchant and Neely, 2023). Input impedance *Z*_*M E*_ is represented with a phenomenological impedance analogy lumped-element model with three parallel impedances where each impedance consists of elements of compliance *K*_*k*_, resistance *R*_*k*_, and mass *M*_*k*_ in series.

To fit the ear canal and middle and inner ear model to our WBT data, we adjust a total of 17 parameters, including 9 middle and inner ear components (*M*_*k*_, *R*_*k*_, *K*_*k*_) and parameters influencing the ear canal transmission matrix components (*A*_*EC*_, *B*_*EC*_, *C*_*EC*_, and *‘D*_*EC*_). The input impedance *Z*_*M E*_ is calculated as the inverse of the sum of the three parallel admittances:

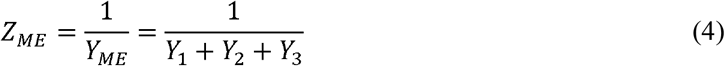

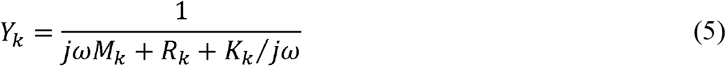

Here, *j* is the imaginary number, *ω = 2πf* is the radian frequency, and *k = 1, 2, 3* denotes the circuit branch number.

The input impedance *Z*_*EC*_ is related to the input impedance *Z*_*ME*_ via the elements of the ABCD ear-canal transmission matrix (Lewis and Neely, 2015):

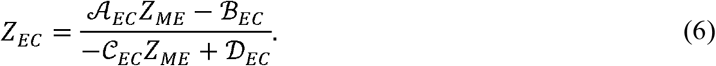

To compute *Z*_*ME*_ from *Z*_*EC*_, we invert the equation:

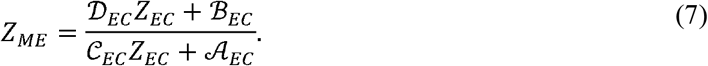

For this assumed relationship, the transmission matrix satisfies the reciprocity condition 𝒜_*EC*_ 𝒟_*EC*_ ℬ_*E*_ 𝒞_*EC*_ *= 1*, and the matrix elements in both Equations (6) and (7) remain the same. In this manner, we can determine the input impedance *Z*_*ME*_ from *Z*_*EC*_, and thus able to focus our efforts on input impedance *Z*_*ME*_ without the varied acoustic influence of the ear-canal.

Model parameters were fitted using a simplex search algorithm, which minimized a cost function. The cost function was a weighted sum of deviations between the measured and modeled ear canal impedance (*Z*_*EC*,*meas*_ and *Z*_*EC*,*model*_) and absorbance (𝔸_*meas*_ and 𝔸_*model*_). For *Z*_*EC*_,_*meas*_ the subscript “meas” denotes values calculated from measured impedance. *Z*_*EC*_,_*model*_ is derived from the fitted middle ear model parameters and Equations (4), (5), and (6). 𝔸_*meas*_ is calculated from *Z*_*EC*_,_*meas*_ and Equations (1), (2), and (3). 𝔸_*model*_ is calculated from *Z*_*EC*_,_*model*_ using the same equations. To ensure numerical stability and realistic parameter estimation, search constraints, such as lower and upper bounds on the model parameters, were incorporated into the cost function as penalty terms.

### Regression Analysis

A multinomial logistic regression model was employed to classify ears into three categories: normal, SCD, and SF. The analysis was implemented using Python’s *scikit-learn* library. Given the high dimensionality of the WBT dataset, where the number of frequency components exceeded the number of subjects, dimensionality reduction was necessary. To address this, principal component analysis (PCA) was applied to transform the WBT and ABG feature space (all 5 individual ABGs) while retaining components that accounted for 90% of the total explained variance. This ensured that the most informative features were preserved while significantly reducing the computational complexity.

The logistic regression models were trained and validated using a stratified k-fold cross-validation approach, ensuring a balanced distribution of the three classes across training and validation sets. Regularization was applied to mitigate overfitting and improve generalizability; for each fold, the model selected whether L1 (sparsity) or L2 (shrinkage) regularization performed better for that feature set. Hyperparameters, including the regularization strength, were optimized to maximize the f1-score, a performance metric that balances precision and recall. In this multiclass case, precision refers to the proportion of predicted cases for a given class that are truly in that class, and recall refers to the proportion of actual cases of that class that are correctly identified. This balanced metric is particularly useful for cases where class imbalance, in which one or more classes are underrepresented, may cause overall classification accuracy to be misleading.

## RESULTS

### Quality of Computational Model Fit

The quality of the analog-circuit model fit was quantified for each ear using the model fit error, which accounts for discrepancies in 𝔸, *Z*_*EC*_ magnitude, and *Z*_*EC*_ phase across all measured frequencies. Lower error indicated a better fit, meaning better agreement between measured and modeled data for a particular ear. **Figure 3** shows a model fit for an ear with SF. This example represents the best fit between modeled and measured data among all ears in the study, with a fit error of 0.004. **Figure 4** shows a model fit for a normal ear. This example represents one of the worst fits across all ears in the study, with a fit error of 0.086. These examples serve as a representation for the range of fits achievable with our computational model.

**Figure 3.**
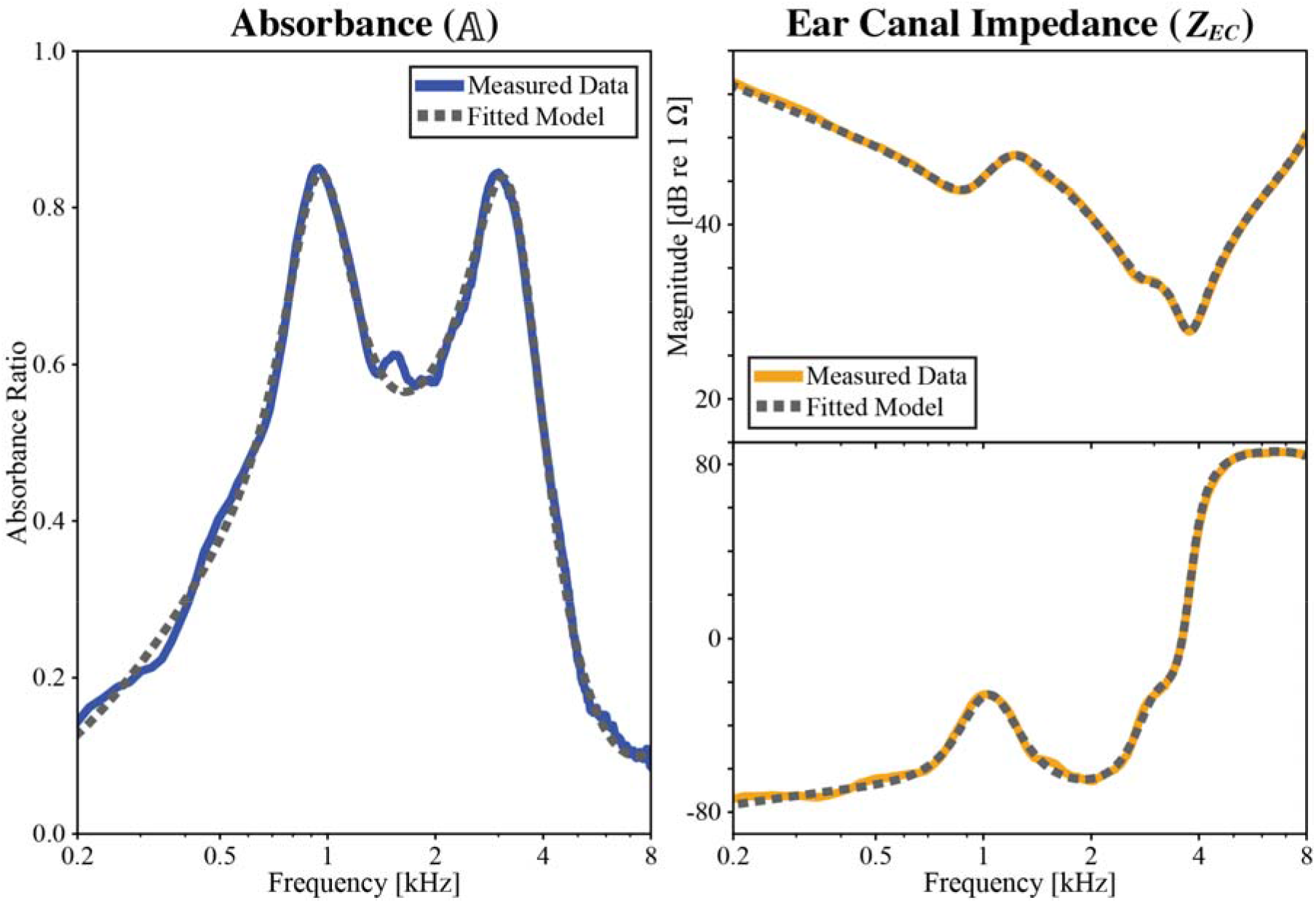
Example of the model fit for an ear with SF. The model fit error for this ear was 0.004, which was one of the best fits with lowest error among the dataset.

**Figure 4.**
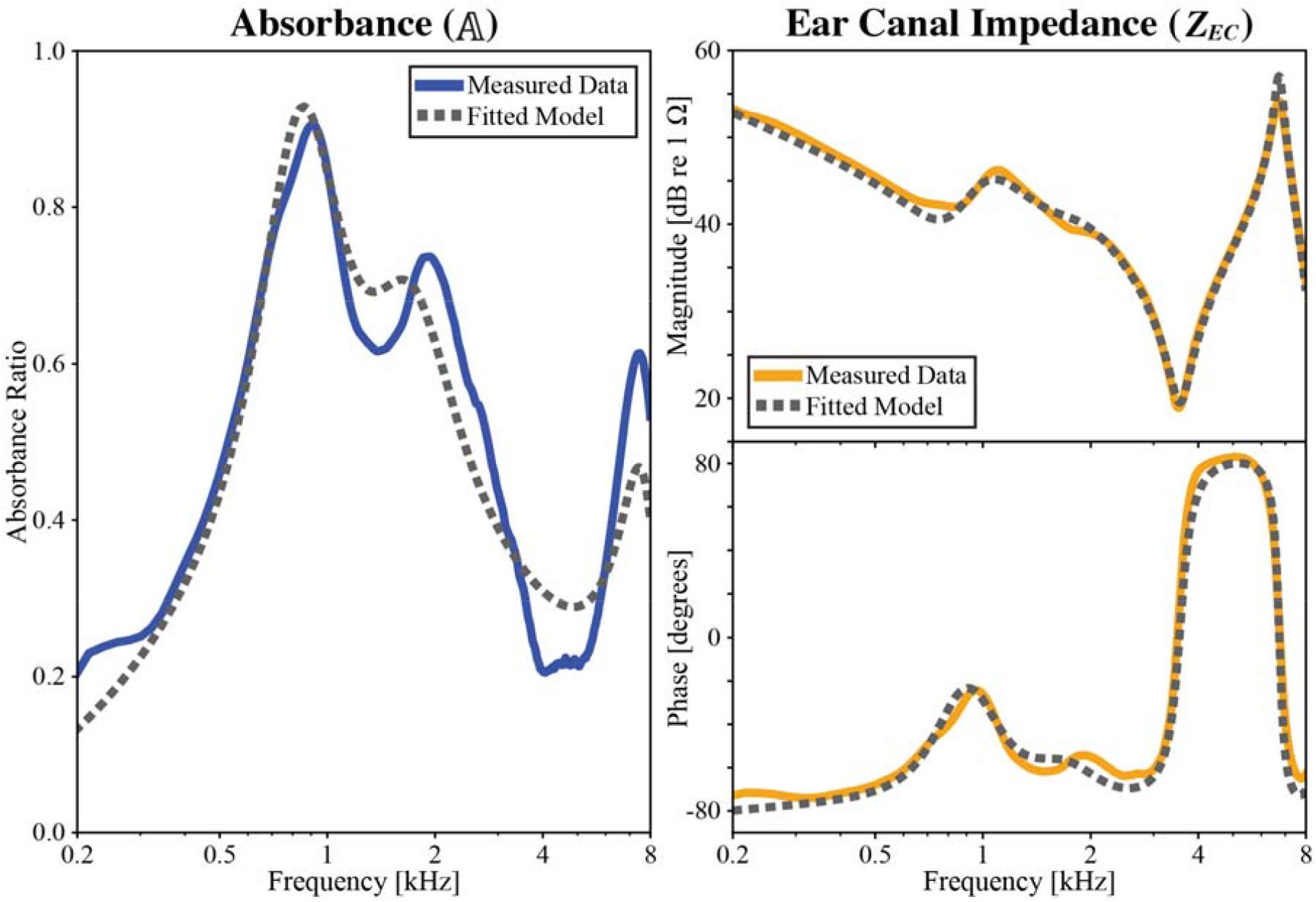
Example of the model fit for a normal ear. The model fit error for this ear was 0.086, which was one of the worst fits with highest error among the dataset.

Figure 5. shows box plots of the model fitting error across analog-circuit models stratified by pathology group. Individual data points are overlaid to illustrate the distribution within each group. To assess whether the quality-of-fit differed between normal and pathological ears, we performed a Kruskal-Wallis test. This test evaluates differences in the rank sums between groups and was appropriate as the model fit error data was not normally distributed. This test revealed a statistically significant difference in model fit error across the three groups (H-statistic = 21.28, p-value = 2.39 × 10^-5^), indicating that at least two of the groups differed significantly from each other. To further investigate, we performed a Dunn’s post hoc test with Bonferroni correction, which accounts for performing multiple comparisons between groups. This test showed a statistically significant difference between the SCD and SF groups (p-value = 1.2 × 10^-5^) where SF had lower fit error than SCD. However, this test showed no statistically significant differences between the normal and SCD groups (p-value = 0.076) or between the normal and SF groups (p-value = 0.114).

### Impedance and Absorbance

Ear canal input impedance is susceptible to variance across measurements even in the same ear due to its sensitivity to probe placement and standing waves within the ear canal. In contrast, middle ear input impedance should remain constant between measurements for a given ear, as it is independent of probe location. **Figure 6** presents representative,, and curves for a normal and pathological SF ear. The examples in **Figure 6** correspond to the same ears displayed in **Figure 3** and **Figure 4**. In both ears, the curves feature a dominating peak between 3 and 4 kHz. This is consistent with expected ear canal resonance frequencies of the effective ear canal length (specifically, the portion that functions as a resonance tube with length between probe tip to tympanic membrane). The peak is absent in the curves, potentially reflecting successful removal of ear canal effects via the analog circuit model fit (**Figure 2**).

**Figure 5.**
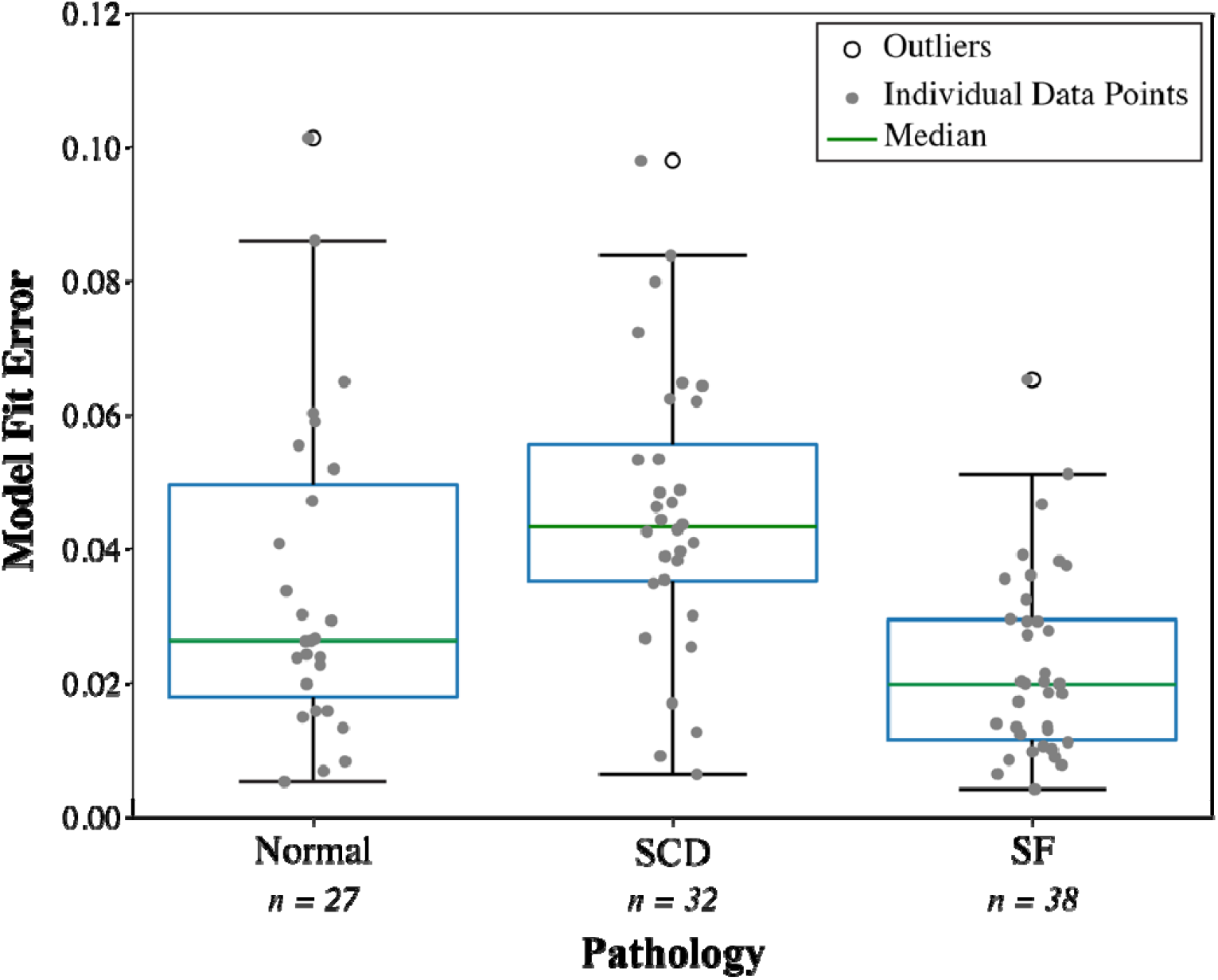
Box plot of model fit error for normal ears (n=27), ears with superior canal dehiscence (SCD, n=32), and ears with stapes fixation (SF, n=38). Individual error values for each ear are shown in gray. The median and interquartile ranges are shown for each group, with outliers marked in black.

**Figure 6.**
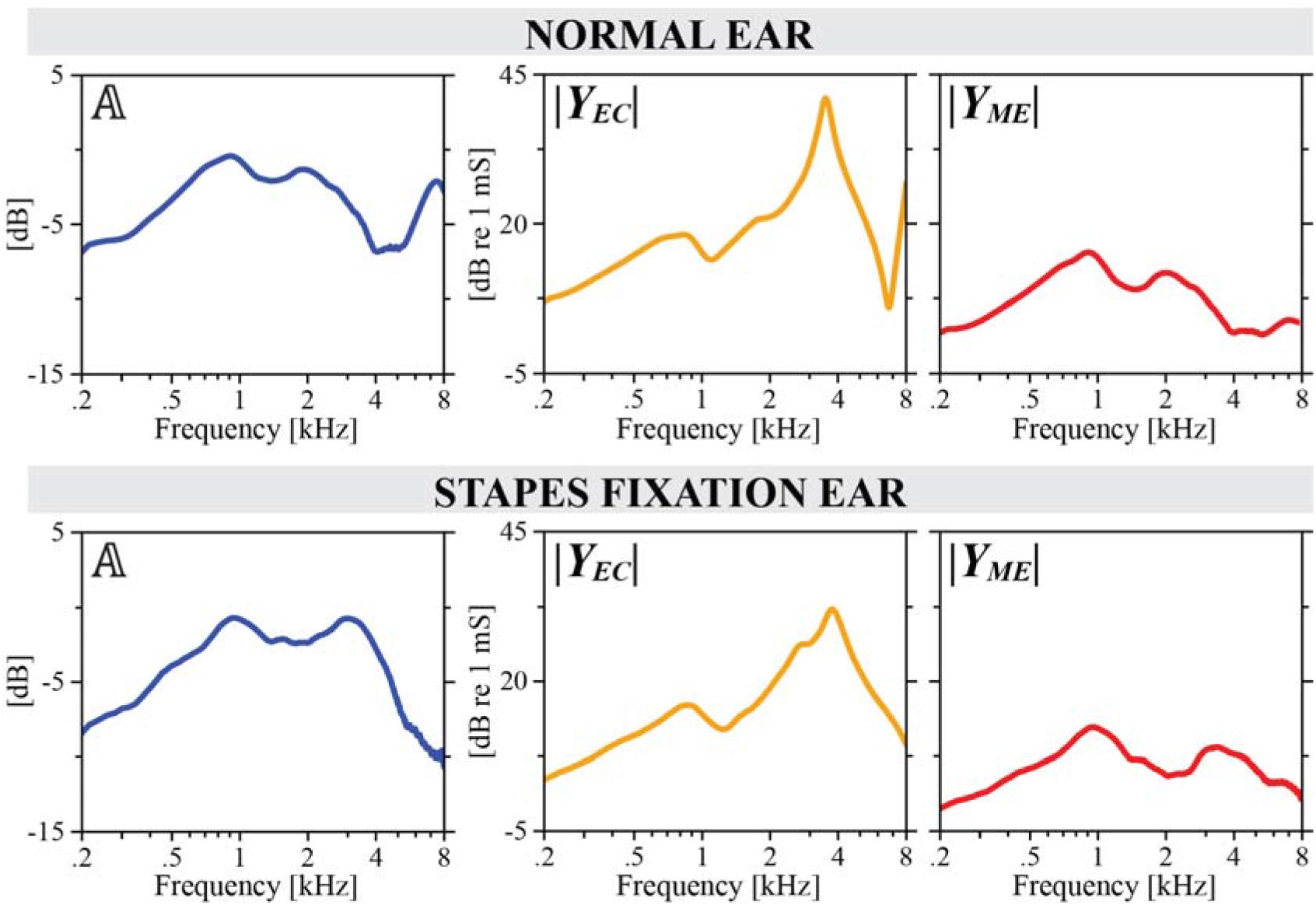
Absorbance (𝔸, blue), ear canal input admittance (|Y_EC_ |, yellow), and middle ear input admittance (|Y_ME_ |, red) curves for a normal ear (top) and pathological ear with SF (bottom). This data corresponds to the same ears as in Figures 3 and 4.

### Classification Performance

Table 1. summarizes the classification performance of the multinomial logistic regression algorithm across different feature sets. Each row represents a different combination of input variables. The percent error for both training and testing data is reported, averaged across five cross-validation folds. While testing error is the primary performance metric, training error is also examined for potential overfitting.

**Table 1.**
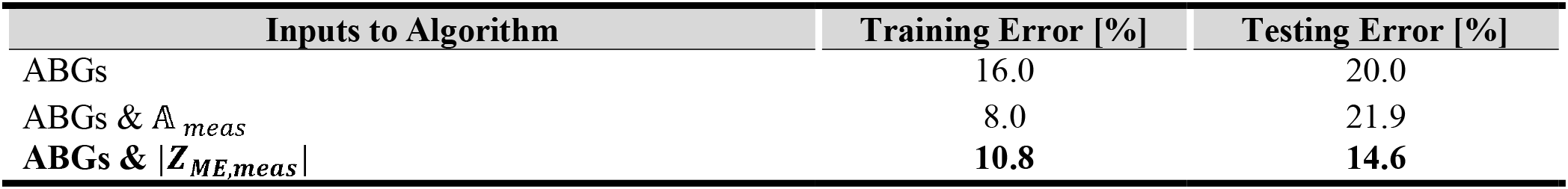
Results of multinomial logistic regression classification algorithm with varying input variables. Training and testing errors are averaged across five stratified cross-validation folds. “ABGs” refers to ABG at all individual frequencies.

Using ABGs (at all frequencies) alone achieved a testing error of 20.0%, demonstrating a reasonable baseline classification performance. We consider this the baseline because the inputs rely solely on ABG features, which are consistently available in clinical evaluations. The similarity between training error (16.0%) and testing error (20.0%) suggests minimal overfitting.

Combining ABGs & absorbance, 𝔸_*meas*_, caused a slight increase in testing error (21.9%) as compared to the case of ABGs alone. The large difference between the training error (8.0%) and the testing error (21.9%) suggests the model may be overfitting the data. Combining ABGs & magnitude of input impedance at the middle ear, |*Z*_*ME*_,_*meas*_|, caused a significant improvement in testing error (14.6%) as compared with the case of ABGs alone or with ABGs & 𝔸_*meas*_. The similarity between training error (10.8%) and testing error (14.6%) suggests minimal overfitting. Combining ABGs & |*Z*_*ME*_,_*meas*_| shows promise for improving classification performance.

**Table 2** displays the overall accuracy and confusion matrices for the classification algorithms with ABGs, ABGs & 𝔸_*meas*_ and ABGs & |*Z*_*ME*_,_*meas*_| as input features. Across each row, the sensitivities and specificities for each group are shown. The sensitivity indicates how well the algorithm correctly identified ears belonging to that class, while specificity indicates how well the algorithm identified ears that did not belong to that class.

**Table 2.**
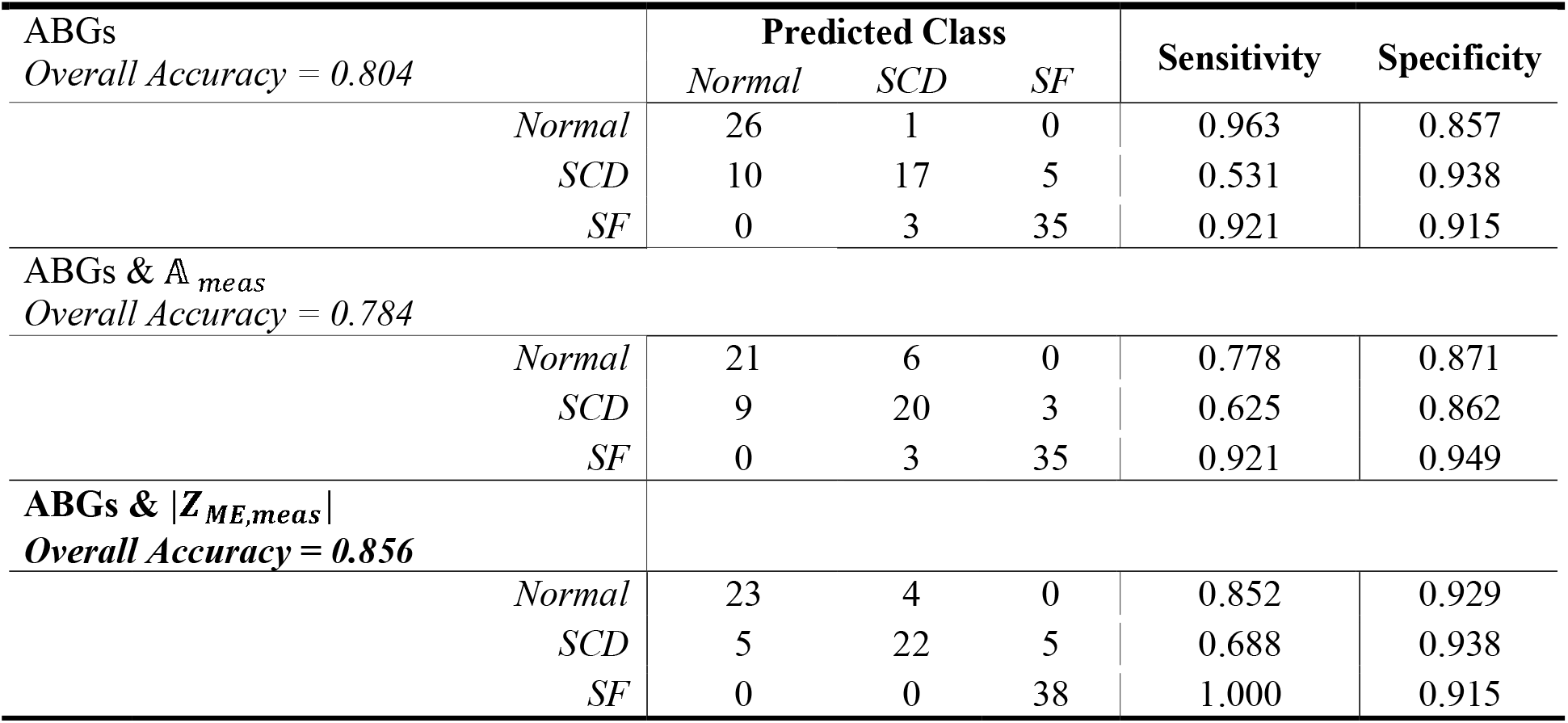
Confusion matrices showing the classification performance with the following algorithm input features: ABGs, ABGs & 𝔸_meas_, and ABGs & |Z_ME_,_meas_ |. Each matrix is a sum of the test performance across a k=5 stratified split, which means each subject in the dataset is accounted for once.

Across the three input feature cases, classification performance varied by class. Using ABGs alone, normal and SF ears were classified with high sensitivity (0.963 and 0.921, respectively), while the sensitivity for SCD ears was substantially lower (0.531). With ABGs & 𝔸 _*meas*_, sensitivity for SCD ears improved to 0.625, while sensitivity for normal ears decreased to 0.778. ABGs & |*Z*_*ME*_,_*meas*_| yielded the highest overall accuracy (0.856) and the highest sensitivity for SCD and SF ears (0.688 and 1.000, respectively). Specificities remained high for all classes across all input feature cases.

## DISCUSSION

In this study, we investigated whether diagnostic classification of normal, SCD, and SF ears could be improved by using measures that better isolate the mechano-acoustic effects of the middle and inner ear. Rather than relying solely on acoustic measurements made at the probe tip, which are influenced by variability in ear canal geometry and probe placement, we applied an analog-circuit model to remove the acoustic contribution of the ear canal and derive the input impedance at the middle ear, *Z*_*ME*_. As *Z*_*ME*_ better reflects the mechanical and acoustic properties of the middle and inner ear, we expected it to improve classification performance relative to absorbance-based approaches.

We evaluated whether incorporating *Z*_*ME*_ into the multinomial logistic regression algorithm would improve diagnostic performance as compared to absorbance,𝔸, used in our previous study (Eberhard et al., 2024). To do this, we implemented an analog-circuit model of the ear (**Figure 2**), which was successfully fitted to measured data (**Figure 3, Figure 4**, and **Figure 5**). From the fitted model, we derived two parameters: input impedance *Z*_*E*_*C*, which appeared to be dominated by standing wave effects (**Figure 6**), and input impedance *Z*_*ME*_. When comparing performance for the best overall classification of normal, SCD, and SF ears, the combination of ABGs & |*Z*_*ME*_,_*meas*_| performed better than the combination of ABGs & 𝔸_*meas*_ (**Table 1**). These results demonstrate that using *Z*_*ME*_ does indeed improve diagnostic performance.

### Limitation of the Computational Model

The analog-circuit model demonstrated a significantly higher model fit error for ears with SCD compared to normal ears and ears with SF (**Figure 5**). This suggests that the model is better suited for characterizing normal ears and ears with SF than for accurately capturing the acoustic characteristics of ears with SCD. Despite these modeling limitations, classification results indicate that the input impedance *Z*_*ME*_ remains a valuable diagnostic feature across all groups tested.

The confusion matrices (**Table 2**) highlight that SCD had the lowest sensitivity across all algorithm inputs (0.531, 0.625, and 0.688, respectively). ABGs & |*Z*_*ME*_,_*meas*_| achieved the highest sensitivity for SCD. Thus, while the analog-circuit model exhibited poorer quality fits for SCD than normal and SF ears, using the model to derive middle ear impedance still resulted in improved algorithm performance. This suggests that SCD may be inherently more challenging to diagnose using ABGs and WBT alone, as compared to normal and SF ears. Since SCD is a lesion of the inner ear, far from the ear canal where WBT measurements are made, changes in inner ear impedance may be more difficult to detect with WBT than those caused by middle ear lesions located closer to the measurement site. In future work, a different or modified model may be needed to more precisely capture the acoustic characteristics of SCD.

### Classification Performance

The best classification performance was achieved with ABGs & |*Z*_*ME*_,_*meas*_|, yielding a 14.6% test error (**Table 1**). This aligns with our expectation that removing the variable acoustic effects of the ear canal would produce a metric, *Z*_*ME*_, that better reflects the middle and inner ear properties affected by SF and SCD than 𝔸_*meas*_. These results highlight the strong predictive power of combining traditional audiometric measure ABGs, with WBT-derived impedance feature *Z*_*ME*_.

It is important to acknowledge that the test errors reported in this study are higher than those reported in Eberhard et al. (2024). The normal subjects included in that study did not all undergo bone conduction testing and, as a result, the ABGs for these subjects were assumed to be zero. This was a limitation of the study, as the machine learning algorithm was able to use the pattern of all-zero ABGs to identify normal subjects, artificially improving the classification performance. For this study, we only included subjects with both air and bone conduction thresholds available.

Additionally, the reported percent errors, sensitivities, and specificities in this study should be interpreted as preliminary. Due to the small dataset size, we used five-fold cross-validation for training and testing the algorithms. This approach means the model was not tested on a fully independent dataset, increasing the risk of overfitting. Consequently, these performance metrics should be viewed primarily as a means of comparing algorithms rather than as indicators of their true clinical performance. Future studies with larger WBT datasets and more advanced machine learning methods will allow for a more robust evaluation.

### Clinical Implications

The findings of this study suggest that incorporating *Z*_*ME*_ into diagnostic algorithms improves the classification of SCD and SF as compared to using 𝔸. The ability to distinguish between different ear pathologies more accurately may reduce reliance on CT scans or exploratory middle ear surgeries, which are used for diagnosis but have risks of radiation and surgery. The approach of using ABGs and WBT, both simple tests for initial evaluation, has the potential achieve accurate diagnosis earlier. By enhancing the accuracy of automated classification algorithms, a non-invasive impedance-based approach could provide an objective, accessible, and cost-effective screening tool for varied clinical settings.

### Future Work

While this study demonstrates the potential of impedance-based classification for diagnosing specific middle and inner ear pathologies, several opportunities exist for future research to enhance the robustness, accuracy, and clinical applicability of our approach.

First, expanding the dataset to include a larger number of pathological cases, as well as additional pathologies, such as ossicular discontinuity and malleus fixation. A broader dataset would improve algorithm generalization and allow for a more comprehensive evaluation of the effectiveness of impedance-based classification across different conditions. A broader dataset would also allow for better differentiation between pathologies that are difficult to diagnostically separate.

Additionally, exploring alternative analog circuit models that are more representative of anatomical structures may improve pathology characterization. More physiologically accurate circuit models may enhance the ability to distinguish subtle impedance variations that are critical for accurate classification. This could also improve the interpretability of algorithm results, as fitted parameters of the analog circuit models would correspond to anatomical features of the ear canal, middle ear, and inner ear.

In this study, we used limited information from our measurements. For example, only the |*Z*_*ME*_ | (magnitude of the middle ear input impedance) was used for the multinomial logistic regression algorithm. Incorporating *∠Z*_*ME*_ (phase of the middle ear input impedance) as an additional diagnostic parameter could provide deeper insights into pathologies by capturing phase-related changes in impedance that may distinguish certain conditions more effectively than magnitude-based metrics alone. Additionally, our study was limited to only tympanic peak pressure (TPP) static pressure. In the future, looking at WBT at other static pressures, such as static pressures close to TPP or large positive or negative pressures, may provide important features to better classify pathologies. Other information, such as additional audiometric measures (e.g. stapedial reflex testing) or symptom-based reporting (e.g. autophony) are often used clinically and may also provide complementary information that could be incorporated as features in our automated diagnostic algorithm.

By exploring these aspects, our goal is to improve the classification algorithm’s robustness, clinical reliability, and diagnostic precision, ultimately increasing its value as a noninvasive and efficient tool for initial diagnosis.

## Data Availability

All data produced in the present study are available upon reasonable request to the authors.

## Notes

### Competing Interest Statement

The authors have declared no competing interest.

### Funding Statement

This work was supported by the National Institutes of Health (NIH) National Institute on Deafness and Other Communication Disorders (Grant T32 DC000038). The contents are solely the responsibility of the authors and do not necessarily represent the official views of NIH.

### Author Declarations

IRB of Massachusetts Eye and Ear gave ethical approval for this work

